# The age-dependent immunogenicity after two doses of MVC-COV1901 vaccine

**DOI:** 10.1101/2021.12.12.21267573

**Authors:** Chia En Lien, Yi-Jiun Lin, Yi-Ling Lin, I-Chen Tai, Charles Chen

**Affiliations:** Medigen Vaccine Biologics Corporation, Taipei, Taiwan; Institute of Biomedical Sciences and Biomedical Translation Research Centre, Academia Sinica, Taipei, Taiwan

## Abstract

A post-hoc analysis of the phase 2 data was performed for the SARS-COV-2 subunit protein vaccine MVC-COV1901. Anti-spike IgG, neutralization assays with live virus and pseudovirus were used to demonstrate age-dependent vaccine-induced antibody response to the vaccine. Results showed that an association exists between age and immune responses to the vaccine, providing further support for the need of booster shots, especially for the older age groups.

## Introduction

MVC-COV1901 is a protein subunit COVID-19 vaccine using the prefusion stabilized spike protein S-2P adjuvanted with CpG 1018 and aluminum hydroxide [1]. MVC-COV1901 has shown favorable safety profile and robust immunogenicity in the phase 2 clinical trial, and has been approved for use in Taiwan since August 2021 [1]. However, how age impacts the immunogenicity elicited by MVC-COV1901 was unknown. In this study, we described the association between age and the antibody titers in 903 healthy adults from 20 to 87 years of age 28 days after the first or second dose in a post-hoc analysis of our phase 2 study.

## Methods

The age-dependent immunogenicity elicited by MVC-COV1901 was observed from the results of three assays, live-virus SARS-CoV-2 neutralization assay, pseudovirus neutralization assay, and the binding of antibodies against SARS-CoV-2 spike. All of the assays followed the same protocols as in the original phase 2 study [1]. Associations between age and NT50 for the Wuhan ancestral strain and the variants were determined by linear regression in Graphpad Prism 8.0.

## Results

The live-virus neutralization assay showed a trend of decreasing immune responses to the vaccine as age increases (Fig 1A). The youngest group of participants (20-29 years; N=188) had a GMT of 914.7 (95% CI, 834.5 to 1002.5), compared to the oldest group (80-87 years; N=7) which has a GMT level of 315.4 (95% CI, 225.1 to 441.9). This is a 65.5% reduction (P<0.001) in GMT. The binding antibody titers also differed in an age-dependent manner. At 28 days after the first dose, the youngest group of participants (20-29 years; N=188) had a GMT of 748.0 (95% CI, 645.8 to 866.5), and the oldest group (80-87 years; N=7) had a GMT of 145.5 (95% CI, 54.6 to 388.2), showing an 80.5% reduction (P=0.0108). At 28 days after the second dose (corresponding to 57 days after the first dose), the youngest group of participants (20-29 years; N=188) had a GMT of 7541.3 (95% CI, 6897.5 to 8245.2), and the oldest group (80-89 years; N=7) had a GMT of 2300.5 (95% CI, 1216.4 to 4350.8), showing a 69.5% reduction (P=0.0024) (Fig 1B). The age-dependent association appears to be independent of the strain, as neutralization against pseudovirus expressing spike protein of the Wuhan ancestral strain, Alpha, and Gamma variants SARS-CoV2 all showed a similar trend (Fig 2).

**Figure 1.**
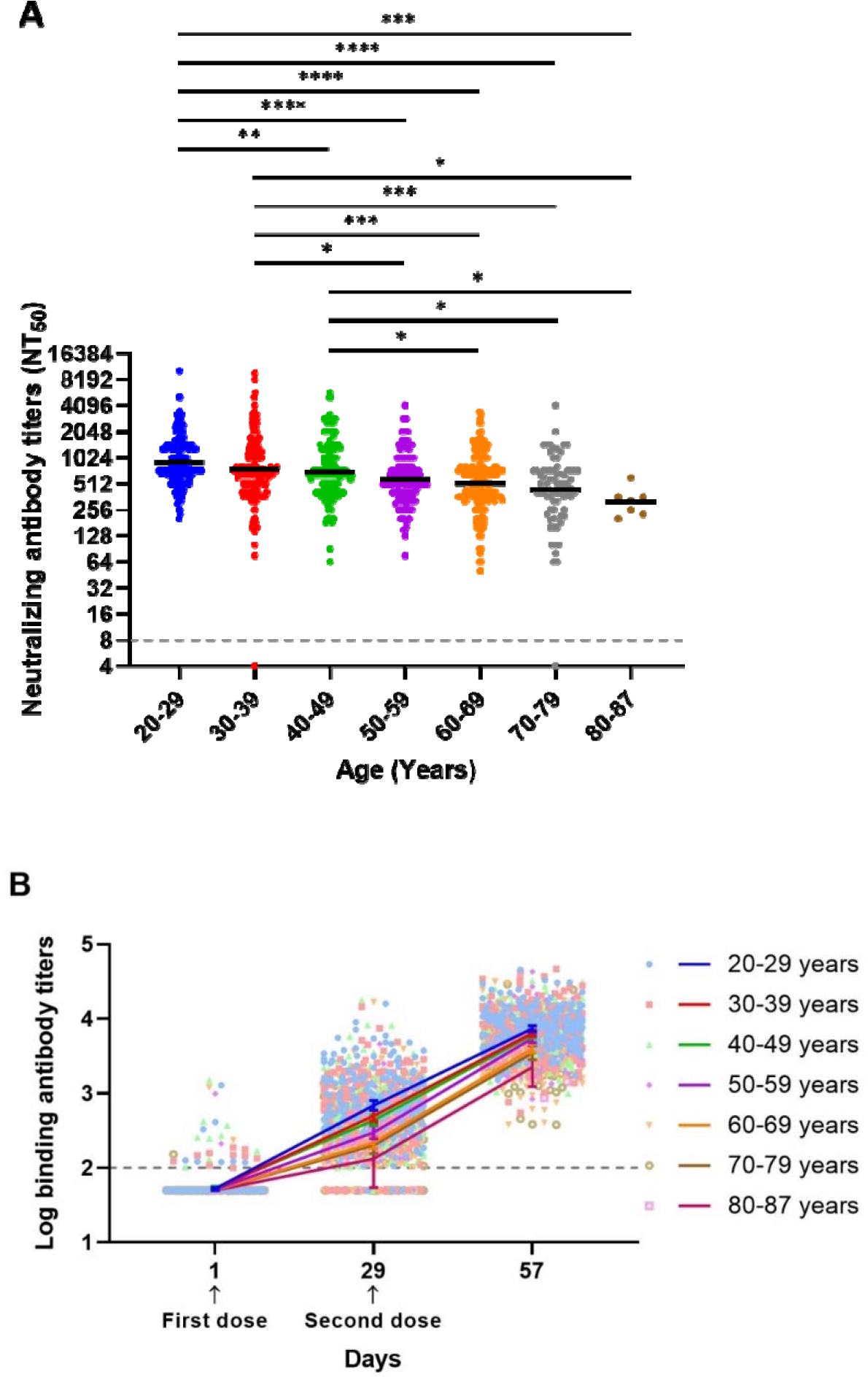
A: Neutralization antibody titers (NT_50_) against SARS-CoV-2 from human subjects vaccinated with MVC-COV1901.Subject age groups are divided by 20-29, 30-39, 40-49, 50-59, 60-69, and 80-87. Each individual neutralization titer is presented by a single-colored dot. Black bar represents the Geometric Mean Titers. Black dotted line represents lower limit of detection. B: Anti-SARS-CoV2 spike protein titers from human subjects vaccinated with MVC-COV1901. Subjects are vaccinated on Days 1 and 29 (black arrows). Each individual titer from the subject is represented by a single-colored dot. Different colors represent different age groups.

**Figure 2:**
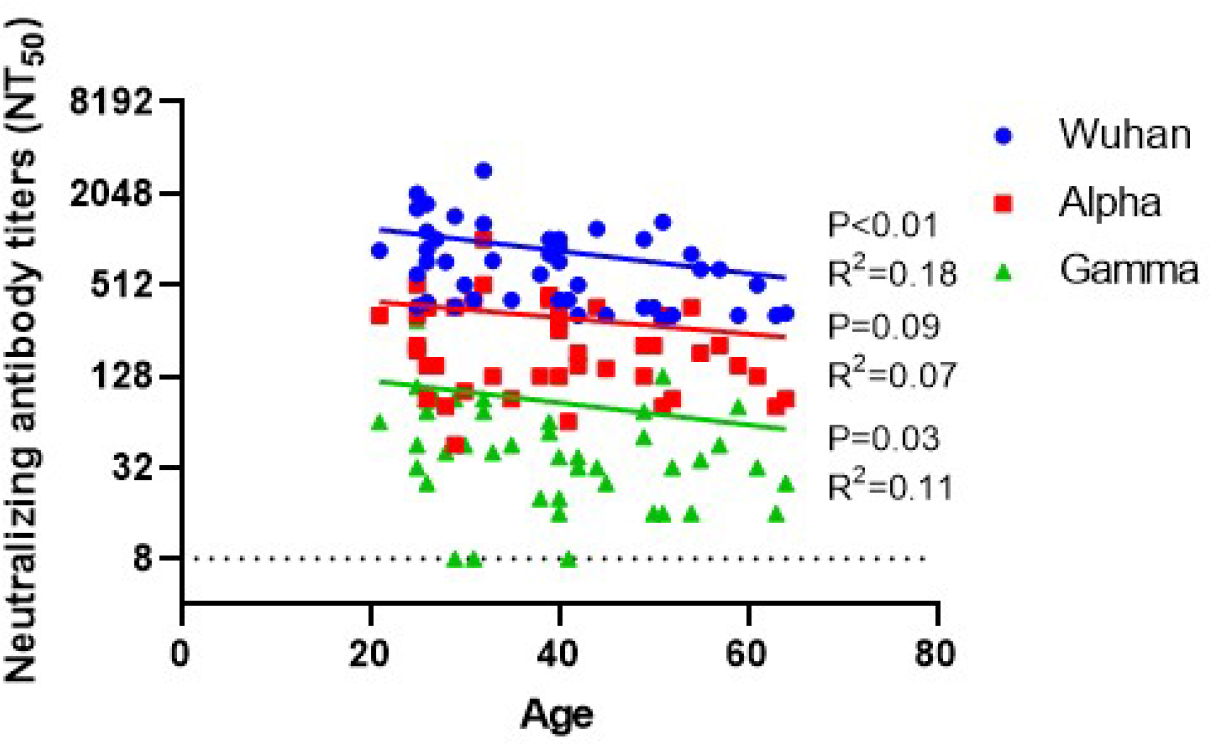
Pseudovirus neutralization titers (NT_50_) against SARS-CoV2 expressing spike protein of the Wuhan ancestral strain, alpha, and gamma variants of SARS-CoV2.

## Discussion

The finding of our post-hoc analysis suggests that protection by the vaccine against SARS-CoV-2 infections is age-dependent since neutralizing antibody levels correlated with age [2]. This is consistent with observations reported by others, that an increase in breakthrough COVID infections was found among older adults [3]. The effect of the age-dependent decrease in immune response appears to be less after the second dose, supporting the positive impact of booster vaccines for older adults. Lastly, the age-dependent decrease in immune response is independent of the virus strains, as similar trends were observed in the ancestral strain, Alpha, and Gamma variants. These reports strengthen the notion that older adults have a lower immune response than younger adults. Second and possibly third dose will likely be important for older adults.

## Data Availability

All data produced in the present study are available upon reasonable request to the authors.

